# Addressing child health inequity through case management of under-five malaria in Nigeria: A model-based extended cost-effectiveness analysis

**DOI:** 10.1101/2021.04.09.21255181

**Authors:** Rishav Raj Dasgupta, Wenhui Mao, Osondu Ogbuoji

**Affiliations:** Duke University, Trinity College of Arts and Sciences, Durham, North Carolina, USA; Center for Policy Impact in Global Health at Duke Global Health Institute, Durham, North Carolina, USA; Duke Margolis Center for Health Policy, Durham, North Carolina, USA

**Author notes:** **Corresponding authors:** Osondu Ogbuoji, 310 Trent Drive, Durham, NC, 27710, Rishav Raj Dasgupta, 310 Trent Drive, Durham, NC, 27710.

## Abstract

**Background:** Under-five malaria in Nigeria remains one of the biggest threats to global child health, accounting for 95,000 annual child deaths. Despite having the highest GDP in Africa, Nigeria’s current health financing system has not succeeded in reducing high out-of-pocket medical expenditure, which discourages care-seeking and use of effective antimalarials in the poorest households. Resultingly, Nigeria has some of the worst indicators of child health equity among low and middle-income countries, stressing the need to evaluate how the benefits of health interventions are distributed across socioeconomic lines.

**Methods:** We developed a decision tree model for case management of under-five malaria in Nigeria and conducted an extended cost-effectiveness analysis of subsidies covering the direct and indirect costs of treatment. We estimated the number of under-five malaria deaths averted, out-of-pocket (OOP) expenditure averted, cases of catastrophic health expenditure (CHE) averted, and cost of implementation.

**Finding:** Fully subsidizing direct medical costs plus a voucher system to cover non-medical and indirect costs with pro-poor increase in treatment coverage would annually avert over 19,000 under-five deaths, US$205.2 million in OOP spending, and 8,600 cases of CHE. Per US$1 million invested, this corresponds to 76 under-five deaths averted, 34 cases of CHE averted, and over US$800,000 in OOP expenditure averted. Due to low current treatment coverage and high disease burden, the health and financial-risk protection benefits would be pro-poor, with the poorest 40% of Nigerians accounting for 72% of all deaths averted, 55% of all OOP expenditure averted, and 74% of all cases of CHE averted. Targeted subsidies to the poor would see significantly greater health and economic benefits per US$1 million invested than broad, non-targeted interventions.

**Conclusion:** Subsidizing case management of under-five malaria for the poorest and most vulnerable children would significantly reduce illness-related impoverishment and child mortality in Nigeria while preserving limited financial resources.

## INTRODUCTION

Despite significant progress in global malaria control over the last two decades, malaria remains one of the leading causes of morbidity and mortality in children under the age of five, who account for two-thirds of the global malaria burden.[1-2] Recent trends also indicate that progress in malaria control is slowing in the highest burden countries.[3] 25% of the global malaria burden is attributable to Nigeria, where malaria annually accounts for an estimated 60% of outpatient hospital visits, 50 million cases, and 100,000 deaths.[4-6] The most vulnerable Nigerians are under-five children, who experience an average of 2-4 episodes of malaria per year and account for as much as 90% of national malaria mortality.[7,8] Moreover, malaria is a leading cause of child death in Nigeria, accounting for as much as 36% of under-five mortality.[9,10]

While prompt and effective treatment of malaria has good clinical outcomes in under-five children, cases where treatment is absent, delayed, or ineffective can become severe and lead to life-threatening complications.[7] Nigeria’s high under-five malaria mortality is largely attributable to a health financing system that leaves many individuals uninsured, resulting in high out-of-pocket (OOP) medical expenditure that discourages care-seeking behavior, especially among the poor **(Box 1)**.[11] Nigeria has one of the lowest rates of care-seeking for suspected cases of under-five malaria in the world, with just under 20% of all under-fives with fever being brought to health facilities for clinical consultation and parasitological testing.[1]

Even when care is sought, the most effective malaria treatments are prohibitively costly and used by few Nigerians.[12,13] Artemisinin-based combination therapy (ACT), the WHO-recommended first line of treatment for uncomplicated malaria, is 98% effective in producing the adequate parasitological and clinical response (APCR) necessary for a child to be cured but is up to twenty times more costly than significantly less effective monotherapies such as chloroquine and sulphadoxine-pyrimethamine.[14,15] On account of high cost to individuals, ACTs are used to treat only 40% of under-five malaria cases in Nigeria.[16] Financial barriers especially hurt the poorest and most vulnerable Nigerians, who use ACT at about half the rate of the richest.[16]

### Box 1

The financial landscape of malaria treatment in Nigeria

In observance of the United Nations Millennium Development Goals (MDGs) for 2000-2015, which aimed in part to eradicate poverty and reduce child mortality, Nigeria’s National Health Insurance Scheme piloted the Free Maternal and Child Health Program (FMCHP) in 2009.[17,18] Among other services, the FMCHP provided free malaria treatment to under-five children brought to public health facilities in 12 out of 36 states.[18,19] Introduction of the FMCHP coincided with national reductions in child mortality, indicating potential efficacy of the program.[18] However, the FMCHP ended in 2015 with the conclusion of the MDGs, as states were unable to sustain necessary funding.[18] As a result, most Nigerian’s currently pay for malaria treatment out of pocket (OOP).[10]

The OOP cost of treating under-five malaria accounts for nearly half of all household medical expense in Nigeria, significantly contributing to catastrophic health expenditure (CHE) in the poorest households.[20] Generally defined as medical expenditure exceeding 10% of annual income, CHE not only leaves individuals unable to pay for future essential health services but often leads to cycles of poverty.[21,22] Globally, CHE pushes nearly 100 million people into poverty per year, exacerbating child health inequalities. Offering financial risk protection, or protection against illness-related impoverishment resulting from CHE, will be essential to make progress toward several of the WHO’s Sustainable Development Goals (SDGs) for 2016-2030.[23]

To that end, scaling up programs like the FMCHP for the provision of effective malaria treatment could incentivize care-seeking behavior and increase service use of the most effective therapies, which may have a positive, pro-poor impact on child health in Nigeria.[19,23,24] Such an intervention would contribute progress toward several SDGs, namely SDG 1 (reducing poverty), SDG 3 (ensuring good health and well-being at all ages), and SDG 10 (reducing inequality within societies).[1,24] Financing under-five malaria treatment through government subsidies would also offer protection against a major source of CHE in Nigeria, which will help achieve SDG target 3.8: achieving Universal Health Coverage (UHC) that ensures access to essential health-care services and access to safe, effective, quality and affordable essential medicines for all.[24,25]

Prior studies of countries in Sub-Saharan African have demonstrated that appropriate diagnosis and treatment are cost-effective interventions for case management of under-five malaria, but there are knowledge gaps regarding the cost-effectiveness of such interventions in Nigeria in particular, as well as how equitable such interventions would be.[26] Equity consideration in evidence-informed policymaking will be crucial to improving child health outcomes in Nigeria—with an exceptionally high under-five mortality rate amongst its poorest children (14% vs. 5% for the poorest and richest fifth, respectively), Nigeria has some of the worst examples of child health inequity in the world.[27,28,29]

This study estimates the potential health and economic benefits of publicly financing case management of under-five malaria in Nigeria through the provision of government subsidies. Particularly, we apply extended cost effectiveness analysis (ECEA) to estimate intervention benefits across different socioeconomic groups. To account for constraints in government health budgets, we investigate the effects of multiple financing strategies that offer varying levels of coverage.[30] ECEA is an analytical technique that assesses health interventions across two main dimensions: health gains and financial risk protection afforded.[31] Moreover, ECEA disaggregates the effects of interventions across population strata of interest, allowing policymakers to identify which subgroups within a broader target population would benefit most from an intervention.[31] This study adds new value to existing cost-effectiveness research by for the first time investigating how the health and economic benefits of publicly financing case management of under-five malaria would be distributed across socioeconomic lines in Nigeria, with the goal of informing policymakers in creating targeted, cost-effective interventions that help the most vulnerable people while preserving limited financial resources.[32]

## METHODS

We conducted an extended cost-effectiveness analysis (ECEA) using a decision tree model created with TreeAge Pro Healthcare software, Version 2020 R2. Using demographic and epidemiological data from published literature and unpublished costing data as parameters, we quantified the health and economic effects of three different intervention scenarios over a year of implementation, disaggregated across five wealth quintiles (Q1-Q5 in ascending order of wealth index). The intervention outcomes estimated were the number of under-five deaths averted, OOP expenditure averted, cases of catastrophic health expenditure (CHE) averted, and the cost of implementation.

### Patient and Public Involvement Statement

Patients and the public were not involved in any way in this study.

### Interventions

Case management of an episode of under-five malaria incurs direct medical costs (consultation, appropriate diagnosis, medical supplies, malaria drugs, other drugs), direct non-medical costs (food on the way to the health facility, transportation, other non-medical supplies and services), and indirect costs (income forgone in productive time lost to caring for a sick child).[33,34,35] In Nigeria, non-medical and indirect costs often represent a high proportion of total expense associated with case management, especially for severe cases that require inpatient hospitalization and significant time spent away from work for caregiving.[19,36]

While the FMCHP subsidized the direct medical costs of treatment, off-setting non-medical and indirect costs will more effectively mitigate the economic burden of treating under-five malaria and could further incentivize service use.[19] To consider varying levels of financing capacity, three intervention scenarios were modeled: (1) a 50% subsidy of direct medical costs (50% DMC), (2) a full subsidy of direct medical costs (full DMC), and (3) a full subsidy of direct medical costs in addition to compensating individuals for non-medical and indirect costs through a voucher system (full DMC + NMC + IC). The cost of implementation was estimated from the government perspective.

In general, greater subsidies incentivize better care-seeking behavior. Therefore, the model assumes a higher increase in treatment coverage for the interventions that cover a greater proportion of total costs. For example, a full DMC + NMC + IC subsidy is expected to result in the most substantial increase in care-seeking behavior, as it would essentially pay individuals for their lost productive time.[37] The model also assumes pro-poor increase in treatment coverage in which poorer quintiles have higher hypothetical increase in treatment coverage than wealthier quintiles. This assumption is based on findings that indicate FMCHP clinics may have been serviced more by poorer and more disease-burdened wealth groups.[18]

For the 50% DMC subsidy, we respectively modeled a 2.5, 2, 1.5, 1, and 0.5 percentage point increase in treatment coverage per quintile; for the full DMC subsidy, we respectively modeled a 5, 4, 3, 2, and 1 percentage point increase in treatment coverage per quintile; for the full DMC + NMC + IC subsidy, we respectively modeled a 10, 8, 6, 4, and 2 percent percentage point increase in treatment coverage per quintile.

### Model parameters

Model inputs were disaggregated by wealth quintile **(Table 1)**. The 2018 Nigeria Demographic and Health Survey (DHS) makes certain demographic and epidemiological parameters available on a per quintile basis, such as malaria prevalence, treatment-seeking behavior, and usage of ACTs.[16] However, reviewing the literature yielded few other model inputs disaggregated by wealth index. Therefore, several calculations were made based on empirical evidence to estimate certain non-empirical indices per wealth quintile, such as indirect costs, adherence to ACTs, and cases of under-five malaria. The calculations for estimating select model parameters by quintile are described in detail in the Supporting Information Section I. Parameters were calibrated such that the model accurately estimated the current number of annual under-five malaria deaths in a base case, non-intervention scenario.

**Table 1.**
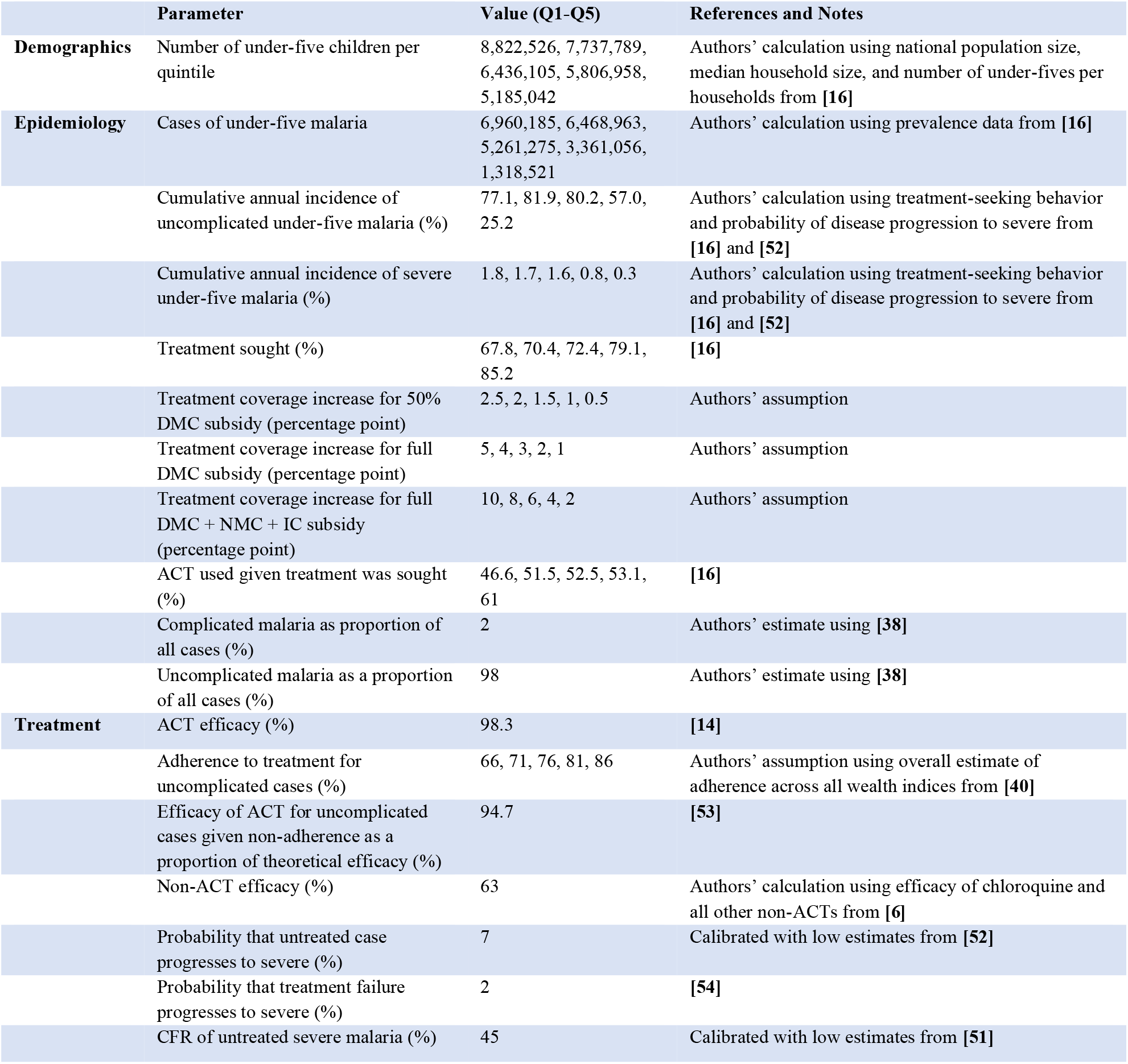
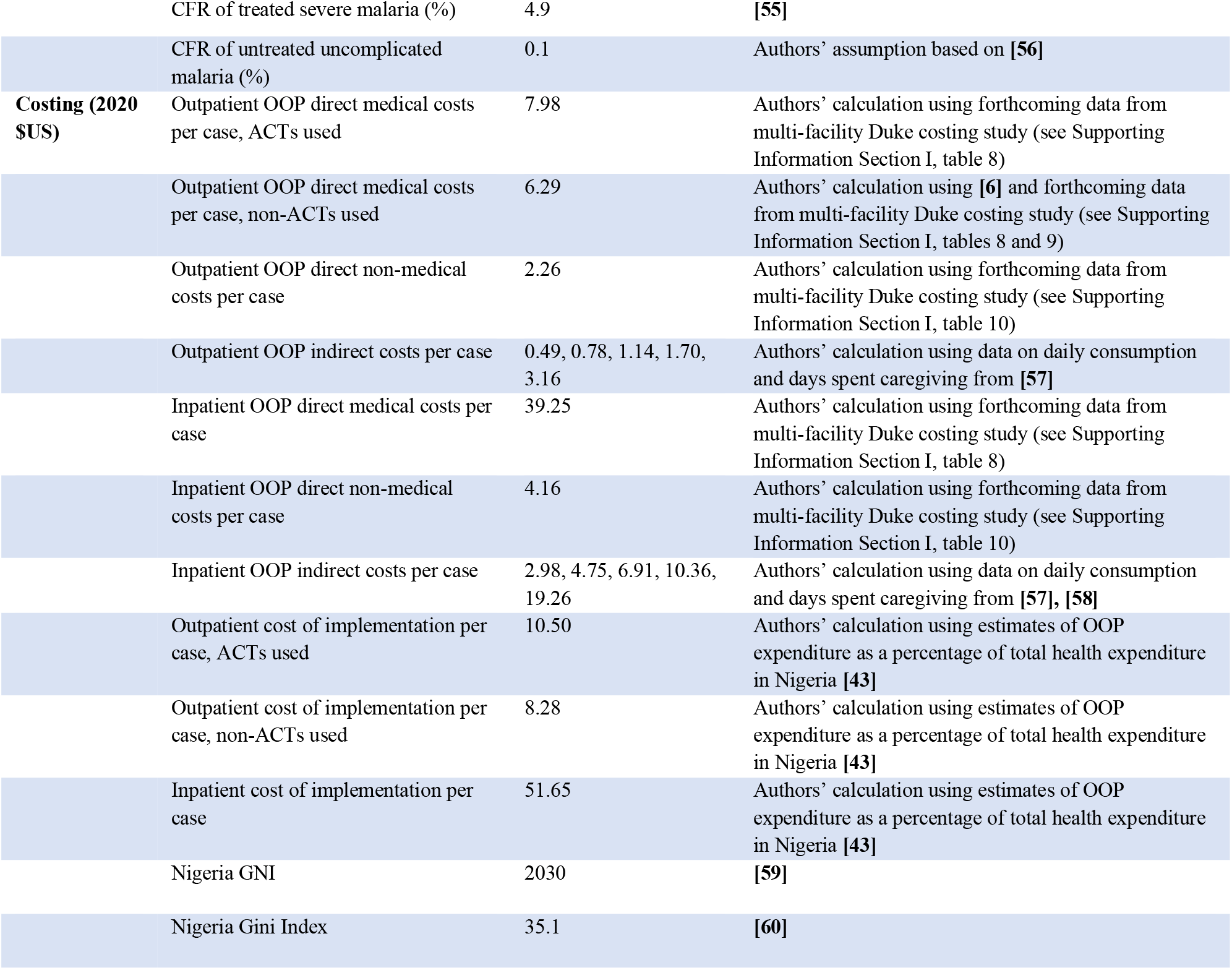
Summary of model parameters.

### Model Flow

TreeAge Pro Healthcare decision analysis software was used to create five decision trees, one for each wealth quintile (Q1-Q5, in ascending order of wealth). Each decision tree is identical in structure, but parameter values differ based on wealth quintile. The entry point for each tree is a case of under-five malaria. The total cases of under-five malaria per quintile were estimated using data from the 2019 World Malaria Report, the U.S. President’s Malaria Initiative Nigeria Malaria Operational Plan for 2020, and the 2018 Nigeria DHS.[1,16,38]

The model’s first chance node splits into clinically treated and untreated cases **(Fig 1A)**. The probability that a case is clinically treated was proxied by the proportion of febrile under-fives for whom treatment is sought, available from the 2018 Nigeria DHS. This approximation was based on the fact that fever in under-fives is a relatively good indicator of malaria in endemic countries.[39] Treated cases split into those treated with ACT and those treated with non-ACTs (e.g., chloroquine, sulphadoxine-pyrimethamine), since ACT is the recommended standard first-line treatment but not always used. Considering that the efficacy of ACT is highly dependent on adherence to the prescribed treatment course, cases treated with ACT split into those both with and without proper adherence.[40]

**Fig 1A:**
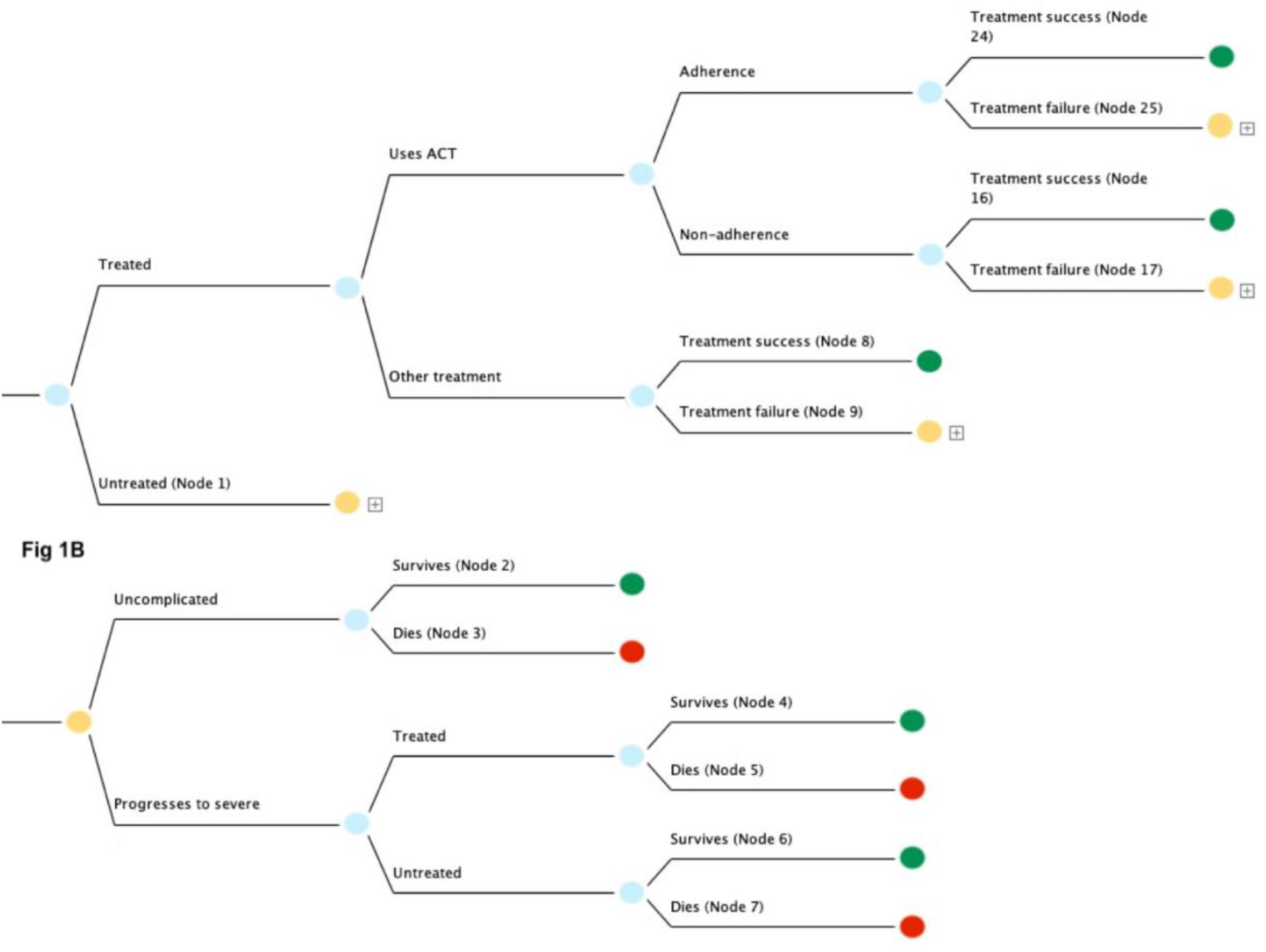
Parent decision tree used to model annual under-five malaria deaths and OOP expenditure in Nigeria. The entry point in the model is a case of under-five malaria. Identical trees were used for each wealth quintile (Q1-Q5) but with different parameters (costs, mortality, and case load). Green terminal nodes are associated with survival. Orange nodes feed into subtrees representing cases that are either untreated (Node 1) or where treatment failure occurs (Nodes 9, 17, and 25). **Fig 1B**: Subtree modelling under-five malaria cases that are either untreated or where treatment failure occurs, representing cases that may progress to severe. Green terminal nodes are associated with survival and red terminal nodes are associated with death.

Both adherent and non-adherent cases were split into treatment success (defined as adequate parasitological and clinical response—i.e., clearance of parasitemia) and treatment failure. Cases not treated with ACT were split directly into treatment success and failure; the effect of adherence on the efficacy of non-ACTs was not considered due to low overall treatment success on account of growing antimalarial drug resistance.[41] Treatment success results in a green terminal node representing survival (nodes 8, 16, and 24).

The orange nodes labeled Node 1, 9, 17, and 25 are associated with cases left untreated or cases where treatment fails; accordingly, these nodes branch into trees that model the progression of disease prognosis to severe **(Fig 1B)**. The trees emanating from nodes 1, 9, 17 and 25 are identical in structure but have parametric differences according to the specific therapeutic scenario, discussed in more detail in Supporting Information Section II. Node 1 is displayed in **Fig 1B**, initially splitting into cases that remain uncomplicated or progress to severe. Uncomplicated cases split into survival and death, respectively resulting in terminal nodes 2 (green) and 3 (red). Severe cases split into cases that are treated and untreated. It was assumed that the probability of seeking treatment for cases that were initially untreated but progressed to severe was the same as the probability of seeking treatment for uncomplicated cases (i.e., treatment of febrile under-fives). Treated and untreated cases each split into survival and death, resulting in terminal nodes 4, 5, 6 and 7.

Using the TreeAge cost-effectiveness setting, cost and effectiveness values were assigned at all terminal nodes. Terminal nodes resulting in survival were assigned an effectiveness value of “1”, and terminal nodes resulting in death were assigned an effectiveness value of “0”. Cost values were assigned according to the OOP cost incurred in each scenario in 2020 United States Dollars (USD). Uncomplicated cases were assumed to incur outpatient costs and severe cases were assumed to incur inpatient costs, an assumption commonly made in malaria modelling studies.[22] For example, terminal node 24, which corresponds to uncomplicated cases treated with ACT, was assigned a total case cost that included outpatient direct medical costs, non-medical costs, and indirect costs **(Fig 1A)**. Terminal node 2, corresponding to untreated uncomplicated cases, was assigned no OOP cost **(Fig 1B)**. Terminal node 4, corresponding to cases that were initially untreated but treated upon progressing to severe, was assigned a total case cost that included inpatient direct medical costs, non-medical costs, and indirect costs. Costing assignments for all terminal nodes are discussed further in Supporting Information Section II. For the 50% DMC intervention, direct medical costs were set at 50% of base case values. For the full DMC intervention, direct medical costs were set at 0. For the full DMC + NMC + IC intervention, all costs were set at 0.

Per quintile for each intervention scenario, annual OOP expenditure and surviving cases were generated stochastically with Monte Carlo simulations using the TreeAge microsimulation tool. The number of random walks simulated per quintile was the estimated number of malaria cases per quintile. A total of about 23 million annual under-five cases were simulated, which is corroborated by prior malaria modeling studies in Nigeria and data from the Institute of Health Metrics and Evaluation (IHME) Global Burden of Disease database.[6,42] For intervention scenarios, increased coverage was simulated by increasing the probability that a case was treated, which is the parameter associated with the very first chance node **(Fig 1A)**. OOP expenditure and surviving cases per intervention scenario were compared against the outcomes of a base case scenario in which treatment coverage was set as the status quo and no subsidy was applied to treatment costs. Additionally, a comparison was made of the incremental OOP expenditure averted between each intervention scenario.

Individual cases of catastrophic health expenditure (CHE) were estimated using a simple disease model, described in Supporting Information Section IV.[31] Cases of CHE were calculated using a threshold of 10% of annual per capita income, and CHE attributable to outpatient and inpatient care were estimated separately.

### Cost of Implementation

For each intervention, the government cost of implementation was estimated using the same decision tree that was used to model deaths and OOP expenditure, but costing parameters were changed to reflect government costs.[22] In Nigeria, OOP spending represents 76% of total current health expenditure while government spending represents 24%.[43] The cost of implementing each intervention we modelled was estimated accordingly.

For the full DMC intervention, where the direct medical costs of treatment are fully subsidized, the government cost of implementation per case was estimated by dividing OOP direct medical costs per case by 76%. For the 50% DMC intervention, the government cost of implementation per case was set as 50% of this value. For the full DMC + NMC + IC intervention, the government cost of implementation per case was set as the sum of total direct medical costs per case (i.e., OOP cost divided by 76%) plus OOP non-medical and indirect costs per case.

### Sensitivity analysis

Uncertainty in model outcomes was quantified using univariate sensitivity analysis. Each model parameter was changed to reflect a high value and low value scenario (respectively 20% higher and 20% lower than the base value). The relative impact of uncertainty in parameter values was quantified by taking the percent difference between the model outcomes of high and low value scenarios and base value scenarios. Uncertainty was estimated for total intervention effects across all quintiles, averaged across all intervention scenarios unless otherwise noted in results.

## RESULTS

### Base case

In the base case scenario without intervention, the model estimated a total of 93,734 annual under-five malaria deaths, 8,637 annual individual cases of CHE, and US$205,221,385 in annual OOP expenditure as a result of treating under-five malaria in Nigeria **(Table 2)**. In this base case, the poorest two quintiles accounted for 66% of mortality, 76% of CHE, and 53% of OOP expenditure. Apart from quintile 1, cases of CHE were exclusively attributable to the cost of inpatient hospitalization.

**Table 2:** Base case annual under-five malaria health and economic indices in Nigeria.

|  |  |  |  |
| --- | --- | --- | --- |
| Wealth quintile | Under-five deaths (thousands) | OOP expenditure (millions, US\$) | Cases of CHE (thousands) |
| Q1 | 34.3 | 54.9 | 4.0 |
| Q2 | 27.9 | 54.2 | 2.6 |
| Q3 | 20.6 | 47.3 | 1.7 |
| Q4 | 8.9 | 33.3 | 0.4 |
| Q5 | 2.0 | 15.6 | 0 |
| Total | 93.7 | 205.2 | 8.6 |

### Deaths averted

The 50% DMC, full DMC, and full DMC + NMC + IC subsidies respectively averted a total of about 4,500, 9,300, and 19,000 under-five deaths **(Fig 2)**. Across all intervention scenarios, health benefits were concentrated among the poor, with the poorest two quintiles accounting for 72% of all deaths averted and the richest quintile accounting for 1% of all deaths averted on average (Supporting Information Section VI, Fig 1).

**Fig 2:**
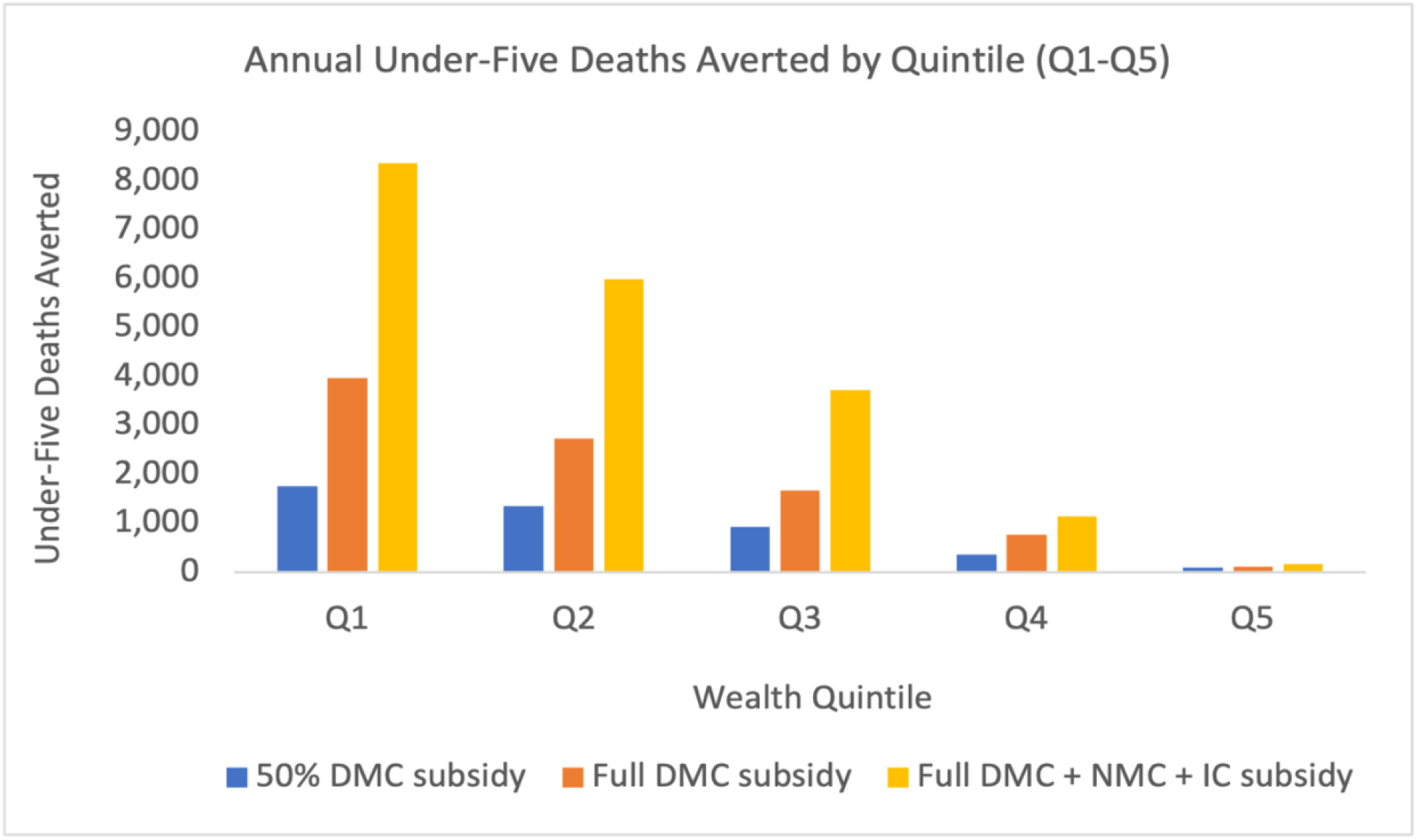
Annual under-five malaria deaths averted by wealth quintile (Q1-Q5) in Nigeria by implementing three different interventions. Deaths averted are concentrated among the poor.

### OOP expenditure averted

The 50% DMC, full DMC, and full DMC + NMC + IC subsidies respectively averted a total of US$70.1 million, US$142.1 million, and US$205.2 million in OOP expenditure across all quintiles **(Fig 3)**. In all intervention scenarios, benefits were concentrated among the poor, with the poorest two quintiles accounting for 55% and the richest quintile for accounting for 7% of total OOP expenditure averted on average (Supporting Information Section VI, Fig 2).

**Fig 3:**
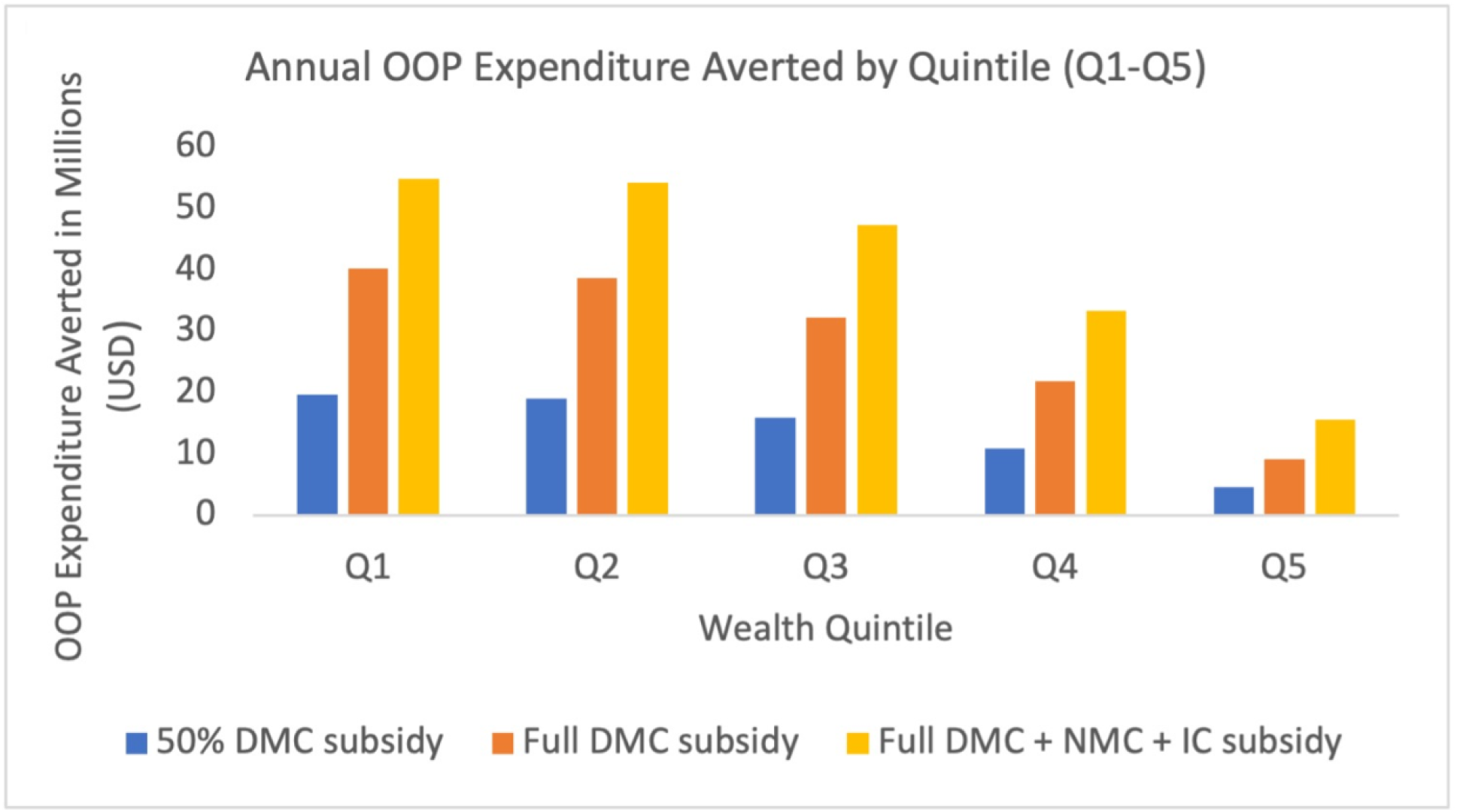
Annual under-five malaria related OOP expenditure averted by wealth quintile (Q1-Q5) in Nigeria by implementing three different interventions. OOP expenditure averted is concentrated among the poor.

In terms of OOP expenditure averted, the incremental benefits across interventions were greater between the 50% DMC and the full DMC subsidies than between the full DMC and full DMC + NMC + IC subsidies. On average across quintiles, the full DMC subsidy resulted in 202% more OOP expenditure averted compared to the 50% DMC subsidy, while the full DMC + NMC + IC subsidy resulted in 49% more OOP expenditure averted compared to the full DMC subsidy **(Figs 4A, 4B)**. The incremental economic benefits of the full DMC subsidy were marginally greater for the poor than the wealthy **(Fig 4A)**, while the incremental economic benefits of the full DMC + NMC + IC subsidy were significantly greater for the wealthy than the poor **(Fig 4B)**.

**Fig 4A:**
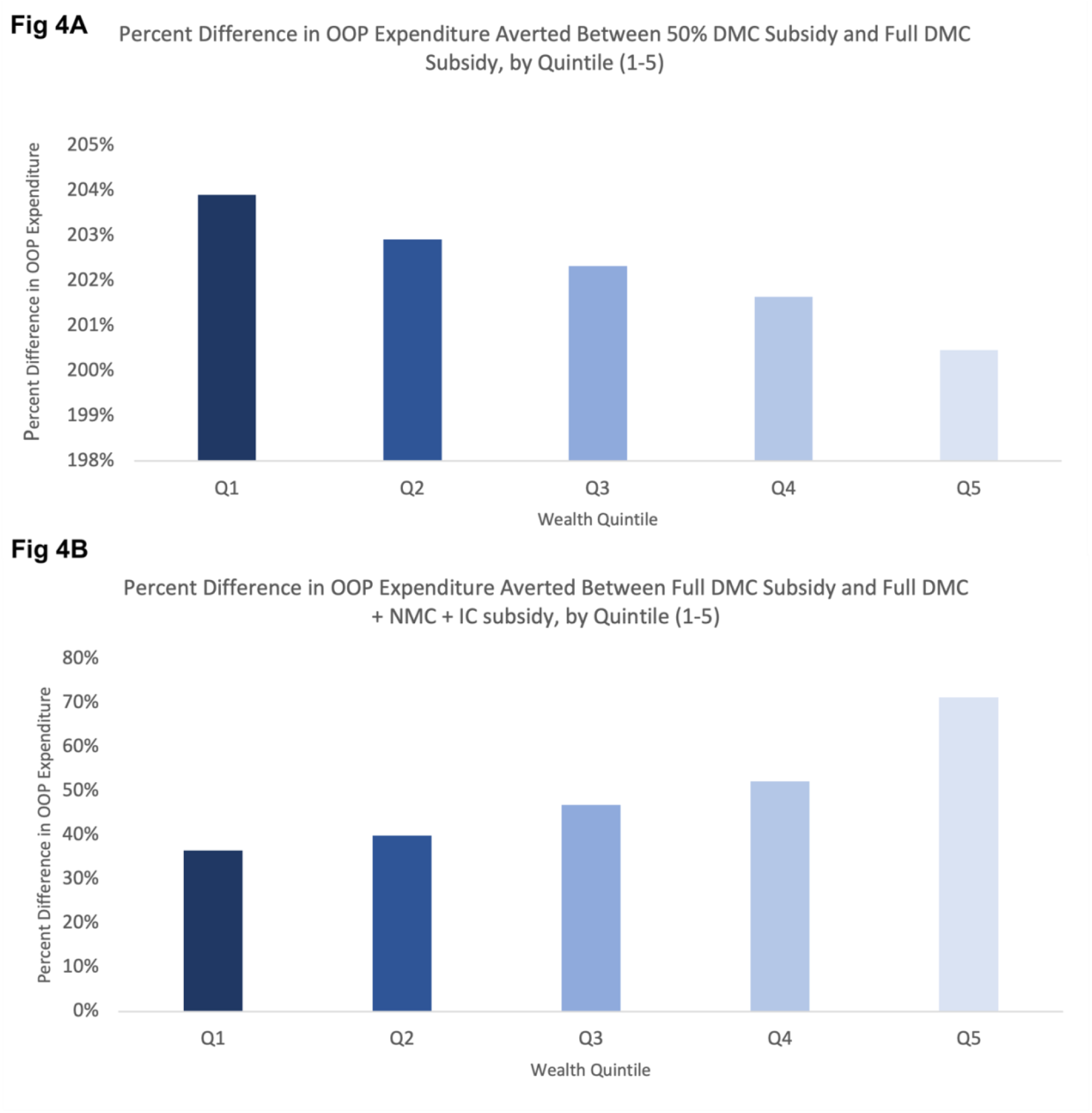
Incremental benefits in OOP expenditure averted between a 50% DMC subsidy and full DMC subsidy by wealth quintile (Q1-Q5). Incremental benefits marginally decrease with wealth. **Fig 4B**: Incremental benefits in OOP expenditure averted between a full DMC subsidy and full DMC + NMC + IC subsidy by wealth quintile (Q1-Q5). Incremental benefits increase with wealth.

### CHE averted

The 50% DMC, full DMC, and full DMC + NMC + IC subsidies respectively averted a total of 7,202, 8,604, and 8,637 annual cases of individual CHE **(Fig 5)**. Across all intervention scenarios, the poorest two quintiles accounted for 74% of all cases of CHE averted on average (Supporting Information Section VI, Fig 3), while quintile 5 experienced no CHE benefits. Only quintiles 1 and 2 experienced incremental benefits between the 50% DMC and the full DMC subsidies, with quintile 1 experiencing greater incremental benefits. Additionally, only quintile 1 experienced incremental benefits between the full DMC subsidy and the full DMC + NMC + IC subsidy, which were marginal.

**Fig 5:**
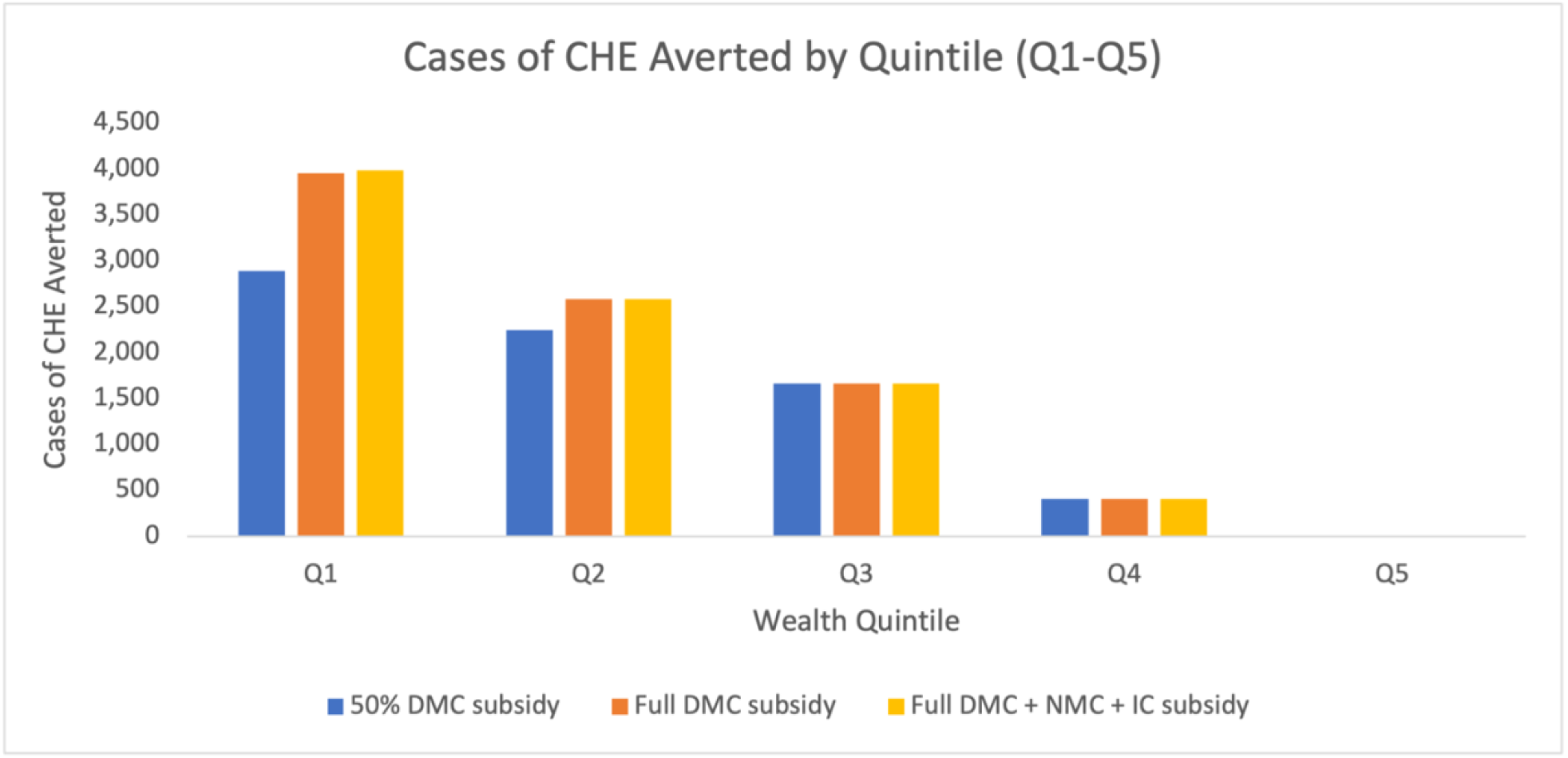
Annual under-five malaria related CHE averted by wealth quintile (Q1-Q5) in Nigeria by implementing three different interventions. CHE averted is concentrated among the poor.

### Cost of implementation

The 50% DMC, full DMC, and full DMC + NMC + IC subsidies would respectively cost the government US$90.5 million, US$179.1 million, and US$254.4 million to implement over one year **(Table 3)**. Across all intervention scenarios, the majority of expenditure would go toward treatment coverage for the poorest Nigerians, with the poorest two quintiles accounting for 56% of total intervention costs on average (Supporting Information Section VI, Fig 4).

**Table 3:** Cost of implementation (in millions, US$)

| **Table 3: Cost of implementation (in millions, US\$)** | | | | | | |
| --- | --- | --- | --- | --- | --- | --- |
| Wealth quintile | Q1 | Q2 | Q3 | Q4 | Q5 | Total |
| 50% DMC subsidy | 25.7 | 24.7 | 20.4 | 13.9 | 5.8 | 90.5 |
| Full DMC subsidy | 51.1 | 48.9 | 40.3 | 27.4 | 11.5 | 179.1 |
| Full DMC + NMC + IC subsidy | 70.5 | 68.2 | 57.3 | 40.1 | 18.3 | 254.4 |

### Benefits per US$1 million invested in each quintile

Per US$1 million invested in each quintile, deaths and cases of CHE averted were greatest among the poor. For all interventions, investing US$1 million into either of the bottom two quintiles would avert more deaths and cases of CHE than investing broadly across all quintiles. **(Tables 4)**. Per US$1 million invested in each quintile, OOP expenditure averted would be relatively equitable in the intervention scenarios that do not subsidize nonmedical and indirect costs. However, for subsidies of nonmedical and indirect costs, wealthy quintiles would avert more OOP expenditure than poor quintiles **(Table 4)**.

**Table 4:** Benefits per US$ 1 million invested in each quintile through targeted subsidies.

|  |  |  |  |  |  |  |  |  |  |
| --- | --- | --- | --- | --- | --- | --- | --- | --- | --- |
| Scenario | 50% DMC subsidy |  |  | Full DMC Subsidy |  |  | Full DMC + NMC + IC Subsidy |  |  |
| Wealth quintile | Deaths Averted | Cases of CHE Averted | OOP expenditure averted (\$US) | Deaths Averted | Cases of CHE Averted | OOP Expenditure Averted (\$US) | Deaths Averted | Cases of CHE Averted | OOP Expenditure Averted (\$US) |
| Q1 | 68 | 112 | 767,068 | 78 | 77 | 786,639 | 118 | 56 | 777,721 |
| Q2 | 55 | 91 | 773,742 | 56 | 53 | 792,114 | 88 | 38 | 794,950 |
| Q3 | 46 | 81 | 778,850 | 42 | 41 | 798,819 | 65 | 29 | 825,721 |
| Q4 | 26 | 29 | 780,029 | 28 | 15 | 798,075 | 28 | 10 | 829,994 |
| Q5 | 17 | 0 | 781,625 | 10 | 0 | 794,492 | 10 | 0 | 851,721 |
| All quintiles (broad subsidy) | 50 | 80 | 774,472 | 52 | 48 | 793,126 | 76 | 34 | 806,708 |

### Sensitivity Analysis

When changed by 20%, the most impactful model parameters on all ECEA outcomes on average were (1) the total number of annual under-five malaria cases and (2) initial treatment coverage, with average effect sizes of 21% and 24%, respectively (Supporting Information Section VII, table 1). In addition, the number of under-five deaths averted was also particularly impacted by (1) the probability that an untreated case progresses to severe and (2) the case fatality rate of untreated severe cases (effect sizes of 19% and 17%, respectively). OOP expenditure averted was more impacted by uncertainty in outpatient costs than inpatient costs, and uncertainty in direct medical costs was more impactful than uncertainty in nonmedical and indirect costs (effect sizes of 5% and 1%, respectively for outpatient and inpatient direct medical costs). The most impactful parameter on cases of CHE averted was inpatient direct medical cost (effect size of 40%). The cost of implementation was substantially more impacted by uncertainty in the costs of outpatient care than inpatient care (effect sizes of 10% and 1%, respectively). A summary of sensitivity analysis results is reported in the Supporting Information Sections VII.

## DISCUSSION

We used extended cost-effectiveness analysis (ECEA) to estimate the health and economic effects of subsidizing case management of under-five malaria in Nigeria. Our model estimates 93,734 annual under-five malaria deaths in a baseline, non-intervention scenario, 66% of which are attributable to the poorest 40% of children **(Table 2)**. Estimated baseline mortality is comparable to figures reported by the WHO Severe Malaria Observatory and the IHME Global Burden of Disease database, which both report about 95,000 annual under-five malaria deaths in Nigeria.[42,44] Our model also estimates that Nigerians spend over US$200 million annually on case management of under-five malaria and that 53% of this cost is incurred by the poorest 40% of Nigerians. This is a sizeable proportion of the reported US$700 million in annual OOP spending on malaria treatment, prevention, and other costs across all ages.[1] Our model estimates that this high OOP expenditure results in over 8,600 annual individual cases of catastrophic health expenditure (CHE), 76% of which are concentrated among the poorest 40% of Nigerians **(Table 2)**. These results emphasize the extent of child health inequity in Nigeria, stressing the need to employ an equity-focused approach to health policymaking that targets the poorest and most underserved populations rather than the mainstream, one-size-fits-all approach that may inadvertently favor the wealthy.[26,29] This is especially important in light of recent plateaus in donor funding of malaria control programs, which may limit the scale of government interventions.[35,45]

Our analysis is one of the first ECEAs of under-five malaria and to our knowledge the first to investigate how the effects of any health intervention in Nigeria would be distributed across socioeconomic lines. A recent ECEA of malaria interventions in Ethiopia found that scaling up coverage of ACTs would afford the greatest health benefits and financial risk protection relative to scaling up use of indoor residual spray, insecticide-treated bed nets, and a hypothetical vaccine.[22] Consistent with our own analysis, this study also found that the health and economic benefits of a treatment subsidy would be concentrated among the poor. Another ECEA found that compared to other interventions in Ethiopia, such as those addressing childhood diarrhea and pneumonia, malaria interventions would see modest benefits, but this is likely due to the relatively low malaria burden in Ethiopia.[46,47] Our analysis builds on the exiting body of ECEA evidence for malaria interventions in Sub-Saharan Africa by investigating different financing strategies that account for nonmedical and indirect costs and expanding geographic scope to Nigeria, the most malaria-burdened country in the world.[1]

In our case management intervention scenarios, we found that larger subsidies would generally result in greater health and economic benefits **(Figs 2**,**3**,**5)**. The full DMC + NMC + IC subsidy, which averted 76 deaths per US$1 million invested broadly across all wealth groups, resulted in substantially greater health benefits than the full DMC and 50% DMC subsidies, which respectively averted 52 and 50 deaths per US$1 million invested across all wealth groups **(Table 4)**. We also found that across intervention scenarios, benefits would be concentrated among the poor, with the poorest 40% of children accounting for 72% of deaths averted, 55% of OOP expenditure averted, and 74% of all cases of CHE averted on average (Supporting Information Section VI, Figs 1-3).

We found that targeted subsidies of the poor would avert significantly more deaths than non-targeted subsidies applied broadly across all wealth groups. For example, in the full DMC + NMC + IC scenario, US$1 million invested into the poorest 20% of Nigerians would avert 118 deaths, while US$1 million invested broadly across all groups would avert 76 deaths, and US$1 million invested into the richest 20% would avert 10 deaths **(Table 4)**. Additionally, we found that while targeted subsidies of direct medical costs would result in relatively equitable OOP savings across wealth groups, subsidies of nonmedical and indirect costs would result in relatively more OOP savings for the wealthy **(Fig 4B, Table 4)**. For example, in the full DMC + NMC +IC scenario, the wealthiest 20% of Nigerians would save about US$850,000 per US$1 million invested while the poorest 20% would save about US$780,000 per US$1 million invested **(Table 4)**. This is likely because higher income individuals incur higher indirect costs on account of productive time lost to caring for sick children—however, these costs are less likely to be catastrophic with increasing wealth.[36] A full DMC + NMC + IC subsidy targeted towards the poor would result in the greatest number of deaths averted and cases of CHE averted while averting OOP expenditure most equitably.

While a full DMC + NMC + IC subsidy would avert US$63.1 million more OOP expenditure than a full DMC subsidy, we found almost no difference in incremental cases of CHE averted between the two scenarios. Resultingly, a broad full DMC + NMC + IC subsidy would avert 34 cases of CHE per US$1 million invested, compared to 48 cases averted in the full DMC scenario and 80 cases averted in the 50% DMC scenario **(Table 4)**. This suggests that CHE related to case management of under-five malaria is largely attributable to the direct medical cost of treatment, which is supported by our sensitivity analysis (Supporting Information Section VII table 1). While non-medical and indirect costs may not be catastrophic, compensating caregivers for these costs may still incentivize care-seeking because individuals in Nigeria often experience multiple different health problems in a given year (acute respiratory infection, pneumonia, HIV/AIDS, etc.), and the cumulative nonmedical and indirect costs could be catastrophic.[48] Targeted subsidies of the poor would avert significantly more cases of CHE than broad subsidies; in the full DMC + NMC + IC scenario, for example, investing $1 million into the poorest 20% of Nigerians would avert 56 cases of CHE, compared to 34 cases averted if invested broadly and 0 cases averted if invested into the richest 20% **(Table 4)**.

Some limitations in our study should be considered when interpreting the results. First, our decision tree model is static, which assumes that the case load of malaria would remain the same across the duration of the intervention course.[49] This assumption does not mirror the realities of transmissible diseases like malaria, but the dynamics of malarial transmission are unlikely to change significantly within a year of implementation, which is the projected time frame of the scenarios we modelled. Furthermore, effective treatment has been shown to interrupt malaria transmission, so this policy could potentially move Nigeria closer to malaria elimination and lower costs in future years.[50,51] Second, our model assumes that case management of uncomplicated malaria incurs outpatient health facility costs, but uncomplicated malaria is often treated in community settings in Nigeria.[16] The likely impact of this assumption is an overestimation of total OOP expenditure averted and total cost of implementation. Estimates of total financial risk protection, however, are relatively unaffected by this assumption because in our model, CHE is most impacted by the cost of severe malaria cases requiring inpatient hospitalization (Supporting Information Section VII, table 1). Third and most significantly, our methodology approximates initial treatment coverage rates as care-seeking behavior for febrile under-fives, a common assumption made in modelling studies of malaria-endemic countries.[22] However, the etiology of under-five fever can also include infection with HIV/AIDS, acute respiratory infection, anemia and pneumonia, so care-seeking for febrile under-fives may not reflect the true coverage rate for malaria treatment.[48] Initial treatment coverage is one of the most influential variables in our model for estimation of all ECEA outcomes (average effect size of 24% when changed by 20%), so this is a key limitation (Supporting Information Section VII, table 1).

Under-five malaria in Nigeria remains one of the biggest challenges to global child health. Stark health inequities between the rich and the poor necessitate the introduction of targeted interventions that benefit the most vulnerable. Targeted subsidization of case management of under-five malaria may be a pro-poor intervention that leads to significant reductions in national under-five mortality and illness-related impoverishment, contributing progress to several of the Sustainable Development Goals including reducing poverty (SDG 1), ensuring good health and well-being at all ages (SDG 3), and reducing inequality within societies (SDG 10). This study provides context for future research that may inform possible policy recommendations. For example, while our study estimates the costs and benefits of subsidizing case management of under-five malaria, a budget impact analysis should be done to determine whether Nigeria can afford such a policy. Furthermore, if case management subsidies are introduced into Nigeria’s Basic Healthcare Provision Fund (BHCPF), an actuarial analysis should be done to understand how the BHCPF would be impacted.

## Data Availability

All data used are freely available online as cited or within the supplemental text.

https://dhsprogram.com/publications/publication-fr359-dhs-final-reports.cfm

## Acknowledgements

The authors would like to thank Ashwini Sunil Deshpande from the Duke Center for Policy Impact in Global Health for assistance in data extraction from the Nigeria Living Standard Survey, Gavin Yamey from the Duke Center for Policy Impact in Global Health for project support, and Megan Knauer from the Duke Margolis Center for Health Policy for assistance with the TreeAge Pro Healthcare software.

