## Supporting Information for "Addressing child health inequity through case management of under-five malaria in Nigeria: A model-based extended cost-effectiveness analysis"

Carolina, USA

^3^ Duke Margolis Center for Health Policy, Durham, North Carolina, USA

**Supporting Information**

I. **Calculations and assumptions for select decision tree model parameters**

**Under-five malaria cases per quintile**: Using data from the 2019 World Malaria Report, the U.S. President’s Malaria Initiative Nigeria Malaria Operational Plan for 2020, and the 2018 Nigeria DHS, the annual number of under-five malaria cases in Nigeria was estimated by multiplying the annual number of global cases by the proportion of all global cases attributable to Nigeria, and then multiplying by the proportion of cases in Nigeria attributable to under-fives.^1,2,3^ Values are shown in table 1:

| **Table 1: Under-five malaria in Nigeria** | |
| --- | --- |
| Variable | Value |
| Global cases | 228,000,000 |
| Proportion attributable to Nigeria | 0.25 |
| Proportion of cases in Nigeria attributable to under-fives | 0.41 |
| **Number of under-five cases in Nigeria** | **23,370,000** |

The number of under-five malaria cases per quintile was then determined by multiplying the total number of under-five cases in Nigeria by the proportion of cases attributable to each quintile. The proportion of cases attributable to each quintile was estimated using prevalence data from the 2018 Nigeria DHS, which reports prevalence per quintile in two samples of children tested using RDTs and microscopy (total of around 20,000 under-fives across both samples)^3^ Prevalence and sample size per quintile are reported in table 2. The calculated total number of cases per quintile and proportion of total cases attributable to each quintile are shown in table 3:

| **Table 2: Prevalence of under-five malaria based on RDT and microscopy** | | | | |
| --- | --- | --- | --- | --- |
| Wealth quintile | Prevalence (RDT group) | Sample size (RDT group) | Prevalence (microscopy) | Sample size (microscopy) |
| Q5 | 0.107 | 2321 | 0.057 | 1731 |
| Q4 | 0.259 | 2377 | 0.147 | 1765 |
| Q3 | 0.386 | 2398 | 0.242 | 1750 |
| Q2 | 0.503 | 2230 | 0.336 | 1572 |
| Q1 | 0.571 | 2115 | 0.384 | 1479 |

| **Table 3: Proportion of under-five malaria cases attributable to each wealth quintile** | | | | | |
| --- | --- | --- | --- | --- | --- |
| Wealth quintile | Number positive (RDT group) | Proportion of total positive cases | Number positive (microscopy group) | Proportion of total positive cases | **Average proportion of total cases** |
| Q5 | 248 | 0.060 | 99 | 0.053 | 0.056 |
| Q4 | 616 | 0.149 | 259 | 0.138 | 0.144 |
| Q3 | 926 | 0.225 | 424 | 0.226 | 0.225 |
| Q2 | 1122 | 0.272 | 528 | 0.281 | 0.277 |
| Q1 | 1208 | 0.293 | 568 | 0.302 | 0.298 |
| Total | 4119 |  | 1,878 |  |  |

While there are large differences in estimated malaria prevalence based on the use of either RDTs or microscopy, the proportion of total cases attributable to each quintile is quite similar across both diagnostic methods. The total number of cases per quintile was then found by multiplying the estimated proportion of cases attributable to each quintile by the total number of cases found from table 1, as shown in table 4:

| **Table 4: Under-five malaria cases per quintile** | |
| --- | --- |
| Wealth quintile | Number of cases |
| Q5 | 1,318,521 |
| Q4 | 3,361,056 |
| Q3 | 5,261,275 |
| Q2 | 6,468,963 |
| Q1 | 6,960,185 |
| Total | 23,370,000 |

**Efficacy of ACT given non-adherence:** the efficacy of ACT (i.e., treatment success resulting in adequate parasitological and clinical response) for uncomplicated cases was found in the literature, which estimated a theoretical APCR of 98.3% for children under the age of five.^4^ However, the efficacy of ACT in practice is dependent on adherence. On account of a dearth of empirical evidence, a within-host modelling study was consulted to determine the effect size of imperfect adherence on ACT efficacy.^5^

The study reported that imperfect adherence results in a treatment failure rate of 9% compared to 4% with perfect adherence, corresponding to an effect size of 0.947 (i.e., the efficacy of ACT with imperfect adherence is 94.7% of the efficacy with perfect adherence), as shown in table 5:

| **Table 5: Effect size of imperfect adherence on ACT efficacy** | |
| --- | --- |
| Variable | Modelled ACT efficacy |
| With adherence | 0.96 |
| With imperfect adherence | 0.91 |
| Ratio of efficacy with imperfect adherence over efficacy with adherence | **0.947** |

**Efficacy of non-ACT treatment*:*** the efficacy of non-ACTs in producing APCR was estimated by taking the average efficacy of chloroquine and all other non-ACT treatments as reported in an earlier malaria modelling study, shown in table 6^6^:

| **Table 6: Efficacy of non-ACTs** | |
| --- | --- |
| Treatment | Efficacy |
| Chloroquine | 0.54 |
| All others | 0.73 |
| **Average** | **0.63** |

**Adherence to ACT:** associations between socioeconomic status and adherence to ACT are documented, though the exact effect size is difficult to measure empirically. Therefore, the value for adherence reported in published literature was assumed to be the adherence for the middle quintile (0.76).^7^ Adherence for successively higher quintiles was assumed to be 5% higher, while adherence for successively lower quintiles was assumed to be 5% lower, as shown in table 7:

| **Table 7: Adherence to ACT based on socioeconomic status** | |
| --- | --- |
| Wealth Quintile | Adherence |
| Q5 | 0.86 |
| Q4 | 0.81 |
| Q3 | 0.76 |
| Q2 | 0.71 |
| Q1 | 0.66 |

**OOP costing:** OOP Direct medical costs per outpatient and inpatient case were calculated by summing the unit costs of malaria services reported in unpublished data from the forthcoming Duke HSDF Costing Study, which provides average direct medical and non-medical costs of pediatric malaria case-management in health facilities across four study sites in Nigeria (Lagos, FCT Abuja, Imo, and Kaduna). Itemized direct medical costs were reported in 2013 Naira and had to be converted to 2020 Naira using a cumulative inflation rate of 1.99; the cost in 2020 Naira was then converted to 2020 USD using a conversion rate of 380 Naira/USD, the average exchange rate for 2020.^8,9^ Direct medical costs per treated case are summarized in table 8:

| **Table 8: Direct medical unit costs for treating under-five malaria using Duke HSDF study** | | | | | | |
| --- | --- | --- | --- | --- | --- | --- |
| Item | Outpatient Costs (2013 Naira) | Outpatient Costs (2020 Naira) | Outpatient Costs (2020 USD) | Inpatient Costs (2013 Naira) | Inpatient Costs (2020 Naira) | Inpatient Costs (2020 USD) |
| Consultation | 677 | 1,350 | 3.55 | 677 | 1,350 | 3.55 |
| Microscopy | 348 | 694 | 1.82 | 348 | 694 | 1.82 |
| Drugs (ACT)* | 498 | 993 | 2.61 | 670 | 1,337 | 3.51 |
| Hospital stay** | 0 | 0 | 0.00 | 5,795 | 11,556 | 30.37 |
| Total | 1,523 | 3,037 | **7.98** | 12,078 | 24,086 | **39.25** |

*The cost of drugs for outpatient cases was calculated by multiplying the unit cost of Artemether/Lumefantrine (ACT) per tab by the recommended treatment prescription for under-fives as per WHO guidelines (6 tabs).^10^ The cost of drugs for inpatient cases was calculated by summing the cost of a course of parenteral Artemether (in lieu of Artesunate, which was not included in the HSDF study) with the unit cost of an ACT treatment course, as per WHO treatment guidelines.^11^

**The total cost of hospital stay for inpatient cases was calculated by multiplying the hospital fee per day by the average number of days spent inpatient (5) for cases of severe under-five malaria, as reported in prior studies^12^

The outpatient direct medical cost for cases treated without ACTs was calculated as $6.29 (2020 USD), estimated using the average cost of non-ACT drugs from data reported in a previous malaria modelling study of Nigeria and then adding this drug cost to other costs of case management (clinical consultation and testing), as reported in the Duke HSDF costing study in table 8.^6^ Calculations of estimated non-ACT drug costs are summarized in table 9:

| **Table 9: Non-ACT drug costs (2020 USD)** | | |
| --- | --- | --- |
| Facility | Chloroquine | Other |
| Private | 0.41 | 1.4 |
| Pharmacy | 0.51 | 1.47 |
| Retail | 0.25 | 1.47 |
| Average | 0.39 | 1.45 |
| **Overall average** | **0.92** | |

The Duke HSDF costing study also reported data on nonmedical costs (transportation to and from the health facility); however, these data are limited in that they represent average costs across a broad range of services. Nonmedical costs were reported from 2019, so inflation rates were not applied, and costs were converted directly to USD, summarized in table 10:

| **Table 10: Nonmedical unit costs of treating under-five malaria using Duke HSDF study** | | | | |
| --- | --- | --- | --- | --- |
| Item | Outpatient (2019 Naira) | Outpatient (2020 USD) | Inpatient (2019 Naira) | Inpatient (2020 USD) |
| Transportation | 446 | 1.17 | 446 | 1.17 |
| Food | 415 | 1.09 | 1137 | 2.99 |
| Total | 861 | **2.26** | 1583 | **4.16** |

Nonmedical costs were assumed to be the same across quintiles due to a lack of empirical data, but this assumption is likely flawed because poorer individuals living in rural areas may incur greater travel time to health facilities than wealthier individuals living in urban areas.

Indirect costs per case per quintile were estimated by multiplying the median daily individual income per quintile by the number of days lost to caregiving for a case of under-five malaria. Outpatient cases were assumed to incur a loss of one productive day, while inpatient cases were assumed to incur a loss of 6.1 productive days on average, as reported in the literature.^13^ Income was estimated by extracting per capita annual consumption per quintile $k$ from Wave 3 of the Nigeria Household Living Standard Survey, 2015-2016, using Stata.^14^ A cumulative inflation rate of 1.45 was applied for the conversion of consumption in 2016 Naira to 2020 Naira. Per capita annual consumption per quintile was divided by 365 to estimate daily per capita income per quintile.

Consumption was used to proxy income because it is generally considered to be a better, more reliably measured metric of well-being in low-and middle-income due to a large informal sector.^15,16^ Calculations for estimating indirect costs per malaria case are summarized in tables 11 and 12:

| **Table 11: Per capita consumption extracted from Nigeria Household Living Standard Survey (2015-2016), Wave 3** | | | | | |
| --- | --- | --- | --- | --- | --- |
| Wealth quintile | Median annual consumption (2016 Naira) | Median annual consumption (2020 Naira) | Median annual consumption (2016 USD) | Median annual consumption (2020 USD) | Median daily consumption (2020 USD) |
| Q5 | 301,481 | 438,644 | 1,507 | 1,153 | **3.16** |
| Q4 | 162,162 | 235,940 | 811 | 620 | **1.70** |
| Q3 | 108,200 | 157,427 | 541 | 414 | **1.13** |
| Q2 | 74,406 | 108,258 | 372 | 285 | **0.78** |
| Q1 | 46,716 | 67,970 | 234 | 179 | **0.49** |

| **Table 12: Indirect unit cost of treating under-five malaria (2020 USD)** | | |
| --- | --- | --- |
| Wealth quintile | Outpatient | Inpatient |
| Q5 | 3.16 | 19.27 |
| Q4 | 1.70 | 10.36 |
| Q3 | 1.13 | 6.91 |
| Q2 | 0.78 | 4.75 |
| Q1 | 0.49 | 2.99 |

II. **Further decision tree specifications**

Node 9 of the parent tree branches into cases treated *without* ACT where treatment failure occurs **(Fig. 1)** (note: color coding differs from diagrams in main text). Nodes 17 and 24 of the parent tree branch into cases treated *with* ACT where treatment failure occurs (given adherence and non-adherence, respectively) **(Fig. 2).** Node 9 is identical in structure to nodes 17 and 24 but differs in cost parameters for each terminal node, reflecting the difference in cost between ACTs and non-ACTs. A summary of all terminal nodes and associated cost assignments is reported in table 1.

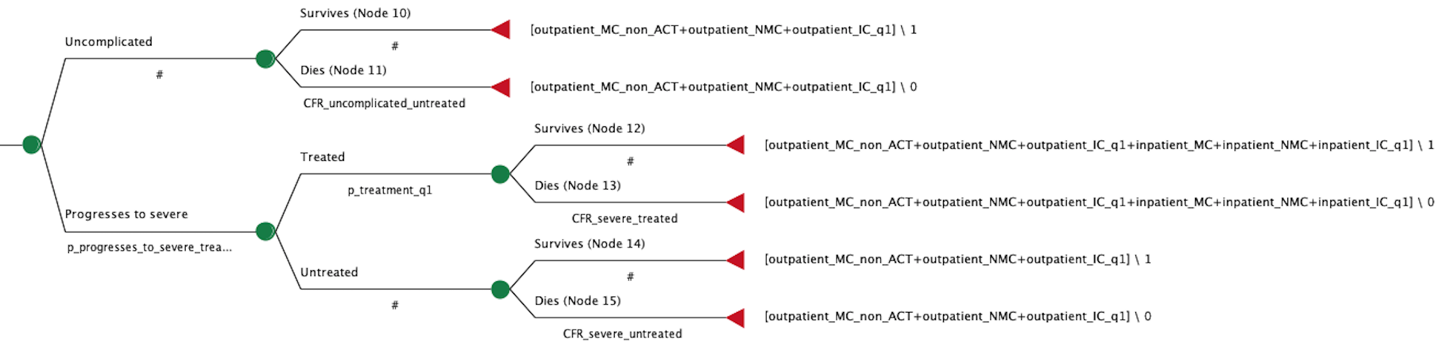
**Fig 1. Node 9**

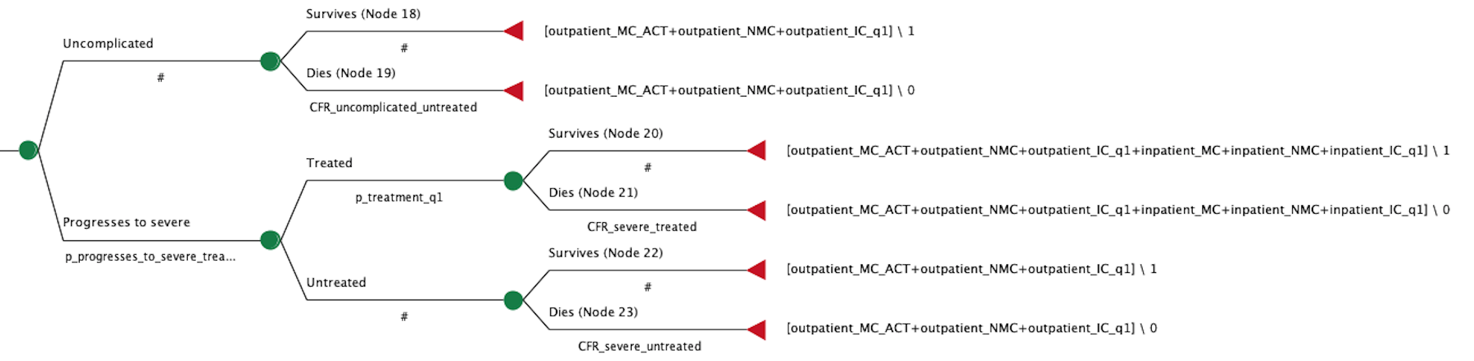
**Fig 2. Nodes 17, 24**

| **Table 1: Summary of costing assignments for terminal nodes** | |
| --- | --- |
| Node | Cost Assignment |
| 2, 3, 6, 7 | 0 |
| 4, 5 | IP direct medical, IP nonmedical, IP indirect |
| 8, 10, 11, 14, 15 | OP (non-ACT) direct medical, OP direct nonmedical, OP indirect |
| 12, 13 | OP (non-ACT) direct medical, OP direct nonmedical, OP indirect, IP direct medical, IP nonmedical, IP indirect |
| 16, 18, 19, 18, 22, 23, 24, 25, 26, 29, 30 | OP (ACT) direct medical, OP direct nonmedical, OP indirect |
| 20, 21, 27, 28 | OP (ACT) direct medical, OP direct nonmedical, OP indirect, IP direct medical, IP nonmedical, IP indirect |

*IP denotes inpatient costs, OP denotes outpatient costs

III. **Extended cost-effectiveness calculations (decision tree model)**

**Deaths averted:** Each intervention scenario resulted in a greater number of surviving cases than the base case. Therefore, the number of under-five malaria deaths averted per quintile $k$ through implementing an intervention were estimated by subtracting the number of surviving cases modeled in the base case from the number of surviving cases modelled in the intervention scenario, as follows:

$$\boldsymbol{D}_{\boldsymbol{a,k}}\boldsymbol{=}\boldsymbol{S}_{\boldsymbol{i,k}}\boldsymbol{-}\boldsymbol{S}_{\boldsymbol{0,k}}$$

Where $D_{a,k}$ are the under-five malaria deaths averted per quintile $k$, $S_{i,k}$ is the number of surviving cases per quintile $k$ in the intervention case, and $S_{0,k}$ is the number of surviving cases per quintile $k$ in the base case.

**OOP expenditure averted:** Each intervention scenario resulted in lower OOP expenditure than the base case. Therefore, the OOP expenditure averted per quintile $k$ through implementing an intervention was estimated by subtracting the OOP expenditure modeled in the intervention scenario from the OOP expenditure modeled in the base case, as follows:

$$\boldsymbol{C}_{\boldsymbol{a,k}}\boldsymbol{=}\boldsymbol{C}_{\boldsymbol{0,k}}\boldsymbol{-}\boldsymbol{C}_{\boldsymbol{i,k}}$$

Where $C_{a,k}$ is the OOP expenditure averted per quintile $k$, $C_{0,k}$ is the OOP expenditure per quintile $k$ in the base case, and $C_{i,k}$ is the OOP expenditure per quintile $k$ in the intervention case.

IV. **Extended cost-effectiveness calculations (simple disease model)**

**Cases of individual CHE averted:** CHE was estimated at the annual level and defined as treatment cost exceeding 10% of annual per capita income, a common threshold of CHE reported in the literature.^18^ The model is adopted from prior ECEA methodology and follows the basic form:

$${\boldsymbol{CHE}_{\boldsymbol{k}}\boldsymbol{=}{\boldsymbol{C}_{\boldsymbol{k}}\boldsymbol{* U}}_{\boldsymbol{k}}\boldsymbol{*P}}_{\boldsymbol{CHE,k}}\boldsymbol{*}\boldsymbol{I}_{\boldsymbol{m,k}}\boldsymbol{*}\boldsymbol{Cov}_{\boldsymbol{k}}$$

Where ${CHE}_{k}$ is the number of individual cases of CHE per quintile $k$, $C_{k}$ is the unit cost per treatment, $U_{k}$ is the number of under-five children per quintile $k$, $P_{CHE,k}$is the proportion of individuals who would be afflicted by treatment-related CHE per quintile $k$, $I_{m,k}$ is the cumulative annual incidence of under-five malaria per quintile $k$, and ${Cov}_{k}$ is the treatment coverage per quintile $k$.^17^ CHE attributable to outpatient and inpatient cases were calculated separately and added together to estimate total CHE.

$U_{k}$, the number of under-five children per quintile, was estimated by dividing the population of Nigeria by the median household size to get the total number of households; this number was then divided by five to get the number of households per quintile; the number of households per quintile was then multiplied by the average number of under-five children per household per quintile to get the total number of children per quintile. Data were available from the 2018 Nigeria DHS, summarized in table 1. In our estimations, the total number of under-fives (about 40 million) represents 20% of the Nigerian population, comparable to estimates of 17% from the United Nations Department of Economic and Social Affairs, Population Division.^18^

| **Table 1: Nigeria household demographics from 2018 DHS** | |
| --- | --- |
| Population of Nigeria | 206,100,000 |
| Median household size | 5.7 |
| Estimated number of households | 36,157,895 |
| Estimated number of households per quintile | 7,231,579 |
| Average number of under-fives per household per quintile (Q1-Q5) | 1.22, 1.07, 0.89, 0.81, 0.72 |
| Estimated number of under-fives per quintile (Q1-Q5) | 8,822,526, 7,737,789, 6,436,105, 5,806,958, 5,185,042 |

$P_{CHE,k}$, the proportion of individuals per quintile $k$ who would be afflicted by CHE at a 10% threshold, was calculated using an inverse gamma distribution based on Nigeria’s GNI, Gini index, and income ranges generated from the Nigeria Household Living Standard Survey, Wave 3 (2015-2016). An inflation rate of 1.45 was applied and conversion to USD was done using a rate of 380 Naira per USD. The upper bounds of income for each quintile are summarized in table 2:

| **Table 2: Upper bounds of per capita income proxied by consumption (from Nigeria Household Living Standard Survey, Wave 3)** | | | |
| --- | --- | --- | --- |
| Wealth quintile | Upper bound consumption (2016 Naira) | Upper bound consumption (2020 Naira) | Upper bound consumption (2020 USD) |
| Q1 | 60,315 | 87,756 | **230.63** |
| Q2 | 89,726 | 130,548 | **343.10** |
| Q3 | 130,943 | 190,517 | **500.70** |
| Q4 | 204,998 | 298,265 | **783.88** |
| Q5 | 2,750,086 | 4,001,279 | **10,515.84** |

$I_{m,k}$, the annual cumulative incidence of under-five malaria per quintile $k$, was estimated separately for uncomplicated and severe cases. $I_{m,k}$ was estimated using treatment coverage data from the 2018 Nigeria DHS as well as a study based on Delphi surveys, which estimates a 7% probability that untreated malaria in under-fives progresses to severe.^19^

Based on these data, the proportion of uncomplicated and severe cases per quintile was estimated by multiplying the proportion of cases left untreated by the probability that an untreated case progresses to severe. The proportion of cases left untreated was assumed to be the proportion of febrile under-fives for whom treatment was not sought. The resulting calculation yielded an overall proportion of 98.2% uncomplicated cases and 1.8% severe cases across all quintiles; these estimates are corroborated by estimates from the 2020 US President’s Malaria Initiative Operational Plan for Nigeria, which reports that about 98.6% of malaria cases in Nigeria are uncomplicated while about 1.4% progress to severe.^2^ Uncomplicated and severe proportions per quintile are summarized in table 3:

| **Table 3: Proportion of total under-five malaria cases that are uncomplicated and severe** | | | | |
| --- | --- | --- | --- | --- |
| Wealth Quintile | Treatment Sought for Fever | Treatment Not Sought for Fever | Proportion of cases that remain uncomplicated | Proportion of cases that progress to severe |
| Q5 | 0.852 | 0.148 | 0.990 | 0.010 |
| Q4 | 0.791 | 0.209 | 0.985 | 0.015 |
| Q3 | 0.724 | 0.276 | 0.981 | 0.019 |
| Q2 | 0.704 | 0.296 | 0.979 | 0.021 |
| Q1 | 0.678 | 0.322 | 0.977 | 0.023 |
| Overall | 0.750 | 0.250 | 0.982 | 0.018 |

Based on the proportions in table 3, the annual number of uncomplicated and severe cases per quintile could be calculated. Cumulative annual incidence was then calculated by dividing the annual number of cases by the number of children per quintile ($U_{k}$), summarized in table 4:

| **Table 4: Cumulative annual incidence of uncomplicated and severe malaria** | | | | | |
| --- | --- | --- | --- | --- | --- |
| Wealth quintile | Total under-five malaria cases | Uncomplicated cases | Severe cases | Uncomplicated incidence | Severe incidence |
| Q5 | 1,318,521 | 1,304,861 | 13,660 | 0.252 | 0.003 |
| Q4 | 3,361,056 | 3,311,884 | 49,172 | 0.570 | 0.008 |
| Q3 | 5,261,275 | 5,159,627 | 101,648 | 0.802 | 0.016 |
| Q2 | 6,468,963 | 6,334,926 | 134,037 | 0.819 | 0.017 |
| Q1 | 6,960,185 | 6,803,302 | 156,883 | 0.771 | 0.018 |
| Overall | 23,370,000 | 22,960,698 | 409,302 | 0.643 | 0.012 |

The proportion of under-five children with malaria using treatment, ${Cov}_{k},$was assumed to be the proportion of febrile under-fives for whom treatment is sought, available from the 2018 Nigeria DHS. ${Cov}_{k}$ was changed based on the scenario being modelled. The cases of CHE averted per intervention were calculated by subtracting the cases of CHE in the intervention scenario from the cases of CHE in the base case, similar to the calculations for deaths averted and OOP expenditure averted.

V. **Tabulation of all ECEA Results for Malaria Treatment Subsidies**

| **Table 1: Under-five Malaria Deaths Averted** | | | | | | |
| --- | --- | --- | --- | --- | --- | --- |
| Wealth Quintile | Q1 | Q2 | Q3 | Q4 | Q5 | Total |
| 50% DMC Subsidy | 1,757 | 1,356 | 930 | 364 | 99 | 4,506 |
| Full DMC Subsidy | 3,979 | 2,741 | 1,677 | 772 | 114 | 9,283 |
| Full DMC + NMC + IC Subsidy | 8,354 | 5,996 | 3,718 | 1,139 | 174 | 19,381 |

| **Table 2: OOP Expenditure Averted (USD)** | | | | | | |
| --- | --- | --- | --- | --- | --- | --- |
| Wealth Quintile | Q1 | Q2 | Q3 | Q4 | Q5 | Total |
| 50% DMC Subsidy | 19,703,036 | 19,086,998 | 15,916,952 | 10,838,104 | 4,540,942 | 70,086,032 |
| Full DMC Subsidy | 40,175,095 | 38,731,953 | 32,204,656 | 21,854,439 | 9,102,832 | 142,068,975 |
| Full DMC + NMC + IC Subsidy | 54,858,701 | 54,205,170 | 47,294,526 | 33,271,449 | 15,591,539 | 205,221,385 |

| **Table 3: Cases of CHE Averted** | | | | | | |
| --- | --- | --- | --- | --- | --- | --- |
| Wealth Quintile | Q1 | Q2 | Q3 | Q4 | Q5 | Total |
| 50% DMC subsidy | 2,885 | 2,248 | 1,665 | 403 | 0 | 7,202 |
| Full DMC subsidy | 3,952 | 2,583 | 1,665 | 403 | 0 | 8,604 |
| Full DMC + NMC + IC subsidy | 3,985 | 2,583 | 1,665 | 403 | 0 | 8,637 |

VI. **Proportional Effects of Malaria Treatment Subsidies by Wealth Quintile**

**Fig 1.	Fig 2.**

Proportion of Total OOP Expenditure Averted by Quintile (Q1-Q5), Average Across Interventions

|  | 28% 27% 23% 16% 7% |
| --- | --- |
| Q1 | 27.7076% |
| Q2 | 26.9698% |
| Q3 | 22.8082% |
| Q4 | 15.6865% |
| Q5 | 6.82795% |

Proportion of Total Deaths Averted by Quintile (Q1-Q5), Average Across Intervention Scenarios

|  | Average |
| --- | --- |
| Q1 | 41.6533% |
| Q2 | 30.1859% |
| Q3 | 19.2961% |
| Q4 | 7.42376% |
| Q5 | 1.44097% |

**Fig 3.	Fig 4.**

Proportion of Total CHE Averted by Quintile (Q1-Q5), Average Across Interventions

|  | Avg of all subsidies |
| --- | --- |
| 1 | 44.0445% |
| 2 | 30.3851% |
| 3 | 20.5827% |
| 4 | 4.98771% |
| 5 | 0% |

Proportion of Total Implementation Cost Attributable to Each Quintile (Q1-Q5), Average Across All Interventions

|  | Avg |
| --- | --- |
| Q1 | 28.6221% |
| Q2 | 27.2487% |
| Q3 | 22.1026% |
| Q4 | 15.4829% |
| Q5 | 6.54373% |

VII. **Summary of sensitivity analysis results**

| **Table 1: Effect of changing model parameters on ECEA outcomes (averaged across intervention scenarios and quintiles)** | | | | | | | | |
| --- | --- | --- | --- | --- | --- | --- | --- | --- |
|  | Under-five deaths averted | | OOP expenditure averted | | Cases of CHE averted | | Cost of implementation | |
| Parameter being changed | 20% higher | 20% lower | 20% higher | 20% lower | 20% higher | 20% lower | 20% higher | 20% lower |
| Cases of malaria | -23% | 23% | -20% | 20% | -19% | 19% | -20% | 20% |
| Initial treatment coverage | 36% | -36% | -14% | 14% | 28% | -28% | -17% | 17% |
| ACT efficacy | -1% | 1% | 0.1% | -0.1% | - | - | - | - |
| Adherence to ACT | 0.2% | -0.2% | 0.1% | -0.1% | - | - | - | - |
| Probability an untreated case progresses to severe | -19% | 19% | -3% | 3% | -19% | 19% | -0.3% | 0.3% |
| CFR (severe, untreated) | -17% | 17% | - | - | - | - | - | - |
| CFR (severe, treated) | -4% | 4% | - | - | - | - | - | - |
| Outpatient OOP direct medical cost (ACT used) | - | - | -5% | 5% | -2% | 2% | - | - |
| Outpatient OOP direct medical cost (non-ACTs used) | - | - | -4% | 4% | -1% | 1% | - | - |
| Outpatient OOP nonmedical cost | - | - | -4% | 4% | -1% | 1% | -1%* | 1%* |
| Outpatient OOP indirect cost | - | - | -1% | -1% | -0.1% | 0.1% | -1%* | -1%* |
| Inpatient OOP direct medical cost | - | - | -0.5% | 0.5% | -40% | 40% | -1% | 1% |
| Inpatient OOP indirect cost | - | - | -0.2% | 0.2% | -3% | 3% | -1%* | 1%* |
| Inpatient OOP nonmedical cost | - | - | -0.2% | 0.2% | -2% | 2% | -0.1%* | 0.1%* |
| Outpatient cost of implementation (ACTs used) | - | - | - | - | - | - | -10% | 10% |
| Outpatient cost of implementation (non-ACTs used) | - | - | - | - | - | - | -10% | 10% |
| Inpatient cost of implementation | - | - | - | - | - | - | -1% | 1% |
| *for full DMC + NMC + IC scenario only | | | | | | | | |
